# A bibliometric analysis of COVID-19 research in Africa

**DOI:** 10.1101/2021.03.15.21253589

**Authors:** Fatuma H Guleid, Robinson Oyando, Evelyn Kabia, Audrey Mumbi, Samuel Akech, Edwine Barasa

**Affiliations:** Health Economics Research Unit, KEMRI-Wellcome Trust Research Programme, Nairobi, Kenya; Health Services Unit, KEMRI-Wellcome Trust Research Programme, Nairobi, Kenya; Centre for Tropical Medicine and Global Health, Nuffield Department of Medicine, University of Oxford, Oxford, UK

## Abstract

**Background:** The ongoing COVID-19 pandemic has led to an unprecedented global research effort to build a body of knowledge that can inform mitigation strategies. We carried out a bibliometric analysis to describe the COVID-19 research output in Africa.

**Methods:** We searched for articles published between 1st December 2019 and 3rd January 2021 from various databases including PubMed, African Journals Online, MedRxiv, BioRxiv, Collabovid, the World Health Organisation global research database and Google for grey literature. Editorial type publications and papers reporting original research done in Africa and were included. Data analysis was done using Microsoft Excel.

**Results:** A total of 1296 articles were retrieved. 46.6% were primary research articles, 48.6% were editorials type articles while 4.6% were secondary research articles. 20.3% articles used the entire continent of Africa as their study setting while South Africa (15.4%) was the most common country focused setting. 90.3% of the articles had at least one African researcher as author, 78.5% had an African researcher as first author, while 63.5% had an African researcher as last author. The University of Cape Town tops the list with the greatest number of first and last authors. Over 13% of the articles were published in MedRxiv and of the studies that declared funding, the Wellcome Trust was the top funding body. The most common research topics include “country preparedness and response” (24.9%) and “the direct and indirect health impacts of the pandemic” (21.6%). However, only 1.0% of articles focus on therapeutics and vaccines.

**Conclusions:** This study sheds light on the contribution of African researchers to COVID-19 research in Africa and highlights Africa’s existing capacity to carry out research that addresses local problems. However, the uneven distribution of research productivity amongst African countries emphasizes the need for increased investment where needed.

## INTRODUCTION

Since its emergence in China in late 2019^1^, Severe Acute Respiratory Syndrome Coronavirus 2 (SARS-CoV-2), the virus that causes COVID-19, has infected over 83 million people and caused over 1.8 million deaths worldwide as of 05^th^ January 2021^2^. SARS-CoV-2 predominantly infects the airways, and disease can range from asymptomatic and mild respiratory infections to severe acute respiratory distress syndrome, with the latter resulting in organ failure in some individuals and eventually leading to death^3-6^. In Africa, where the first case was reported in February 2020, there have been 2.8 million cases and over 68 000 deaths as of 6^th^ January 2021^7^. The pandemic, in addition to health system constraints and the burden of existing communicable diseases, have put considerable pressure on already weak health systems across the continent. In addition, measures put in place to control the spread of the virus^8^ have led to the closure of schools, businesses and social services which have generated significant setbacks to Africa’s heath programs, economy and communities^9-14^.

As COVID-19 is an emerging infectious disease, the research community has responded rapidly to provide insight into how to control the pandemic and in the development of tests, therapeutics, and vaccines. Research on COVID-19 has been predominantly from China, Europe and the USA^15,16^, which is understandable as these regions have experienced more cases and deaths from the pandemic. For reasons that are still uncertain, Africa has experienced fewer cases and deaths in the initial phase of the COVID-19 pandemic compared to other continents. This suggests that local research that considers the local context to inform contextually relevant mitigation strategies and treatment options are necessary.

In this bibliometric analysis, we describe the type of research on COVID-19 that has been done in Africa, the geographical spread of the research, funding sources and contributions of authors from Africa to highlight local capacity for research and research areas that are neglected.

## METHODS

### Data Source

Published papers and grey literature were searched via a topic search (title/abstract) on the following databases: PubMed, African Journals Online, MedRxiv, BioRxiv, Collabovid, the WHO global research database and Google for grey literature. Searches were restricted to those published between 01 December 2019 and 3^rd^ January 2021.

### Eligibility Criteria and Study Selection

Only articles with a focus on COVID-19 in Africa were included. Four reviewers independently performed study selection and data extraction. There were no restrictions on the type of articles that were included. However, only documents in English language were considered for the analysis. Differences of opinion were settled by referral to a fifth review author.

### Search Strategy

The key search words used were those listed for Africa plus keywords pertaining to COVID-19 or SARS-CoV-2 (full search terms are provided in the supplementary file). These search words were used in the title/ abstract fields. Retrieved articles were manually checked for validity of search strategy and articles that were outside the scope and any duplicates were removed.

### Analysis

Descriptive analyses were conducted to evaluate the characteristics and types of articles retrieved using Microsoft excel. This included titles, authors information including, first author and last author, author countries, and affiliations, journal source, funders and funder countries, the research objective, the study setting and keywords. In order to depict relations among keywords, co-occurrence network analysis was done with VOSviewer (Version 1.6.15), a software tool for constructing and visualizing bibliometric networks. VOSviewer was also used to assess co-authorship among all the authors in the bibliography and an evaluation of how many of them were connected within documents authored or co-authored by individuals was conducted.

## RESULTS

### Publication Output

The initial search yielded 6615 studies. After removing duplicates and articles that did not meet the eligibility criteria, we included 1296 studies in our analysis (Figure 1).

**Figure 1:**
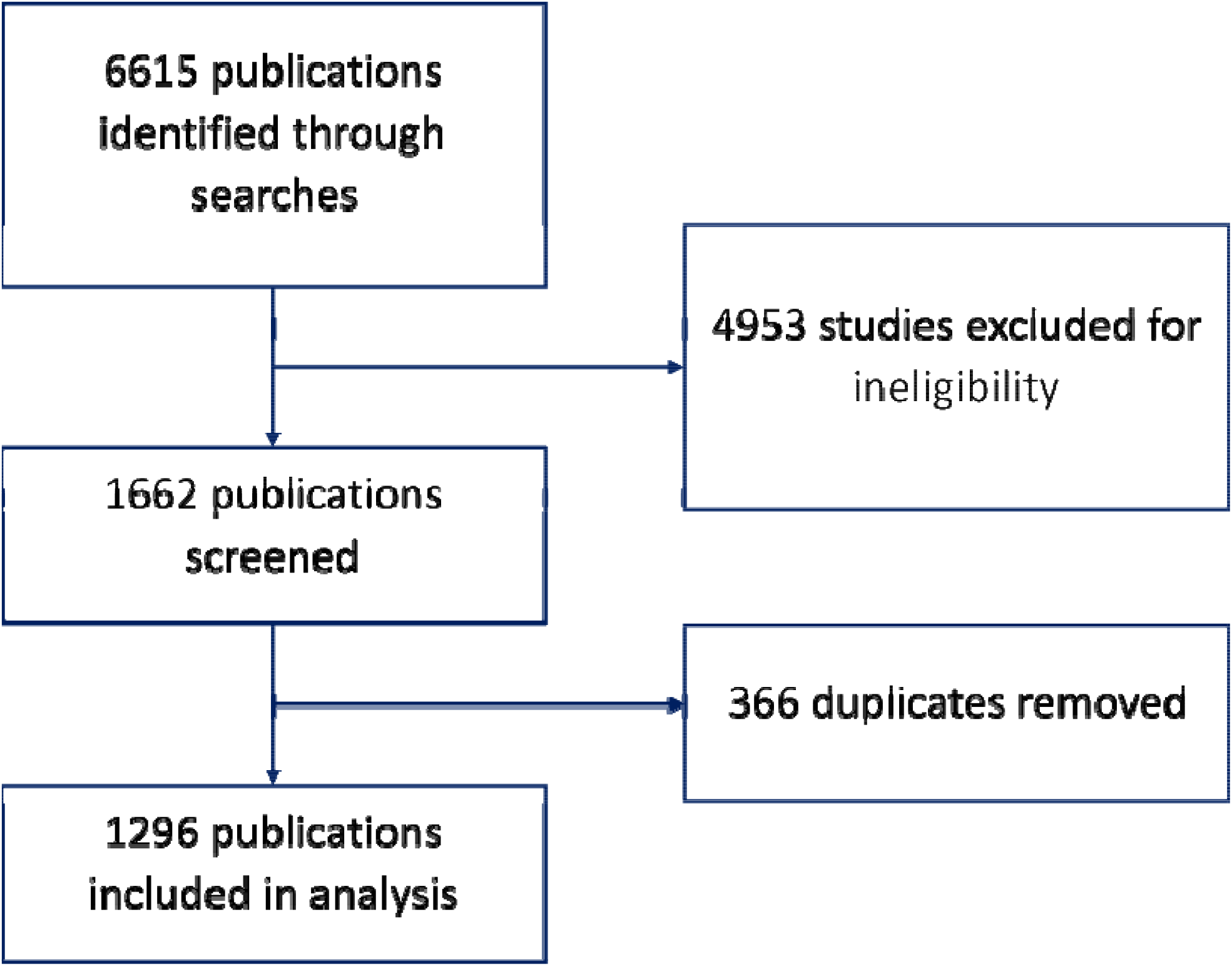
Study flow diagram

Of the 1296 articles reviewed, 630 (48.6%) were non-original research articles (commentary, editorial, perspective pieces etc.), while 606 (46.6%) were primary research articles (research involving the collection of and analysis of primary data) and 60 (4.6%) were secondary research (research involving the analysis of secondary data) articles.

The distribution of countries in which COVID-19 research was conducted is shown in Table 1. 263 (20.3%) articles used the whole of Africa as their study setting while South Africa (15.4%), Nigeria (12.3%), and Ethiopia (6.8%) were the top 3 countries with the highest number of country focused articles (Table 1).

**Table 1:**
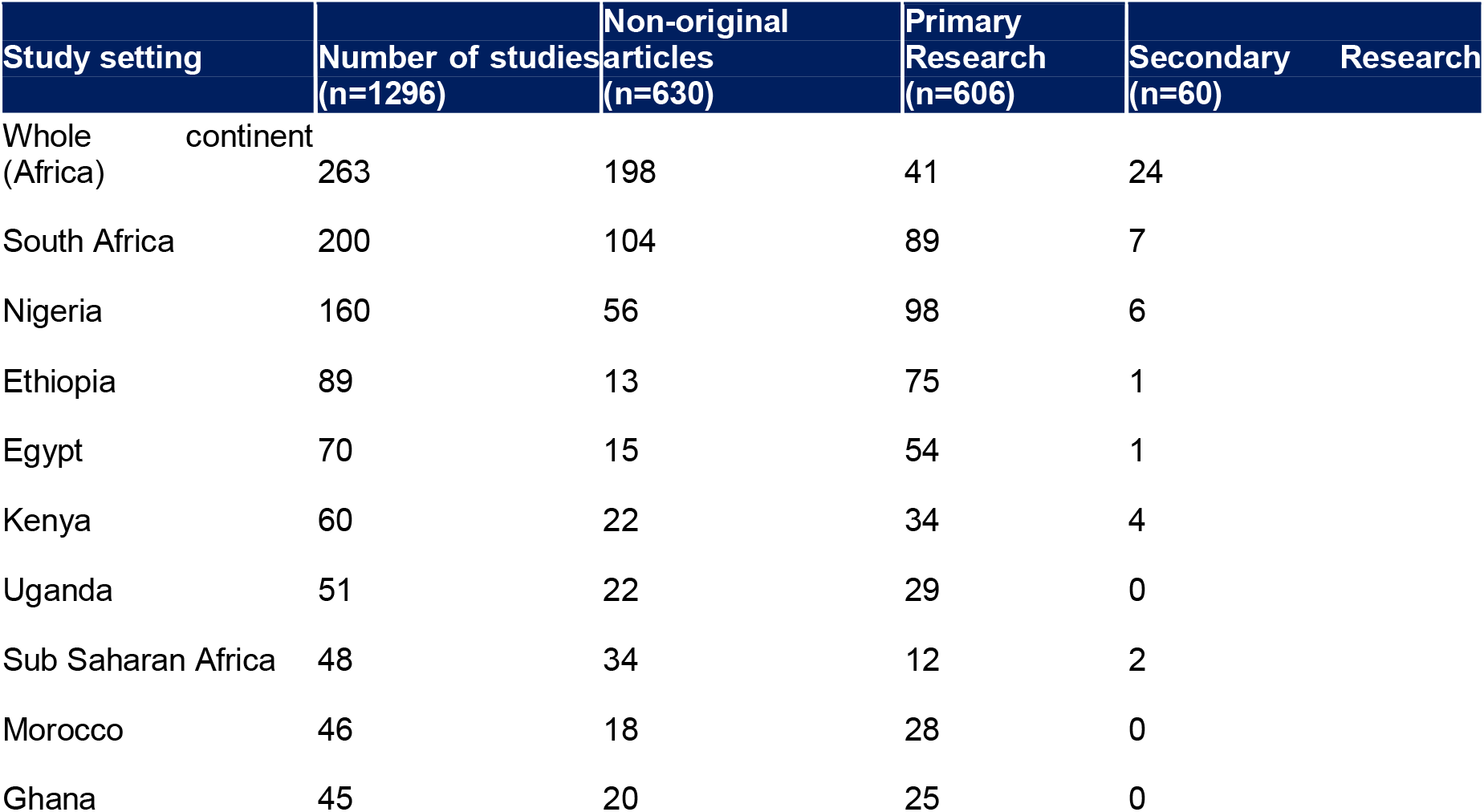
Top 10 study settings in which COVID-19 articles were based on in Africa

| Study setting | Number of studies<br>(n=1296) | Non-original<br>articles<br>(n=630) | Primary<br>Research<br>(n=606) | Secondary Research<br>(n=60) |
| --- | --- | --- | --- | --- |
| Whole continent<br>(Africa) | 263 | 198 | 41 | 24 |
| South Africa | 200 | 104 | 89 | 7 |
| Nigeria | 160 | 56 | 98 | 6 |
| Ethiopia | 89 | 13 | 75 | 1 |
| Egypt | 70 | 15 | 54 | 1 |
| Kenya | 60 | 22 | 34 | 4 |
| Uganda | 51 | 22 | 29 | 0 |
| Sub Saharan Africa | 48 | 34 | 12 | 2 |
| Morocco | 46 | 18 | 28 | 0 |
| Ghana | 45 | 20 | 25 | 0 |

### Authors

A total of 6370 authors were identified. 90.3% of these articles had at least one author affiliated to an African institution. 78.5% of all the articles reviewed had a first author affiliated to an African institution, while 63.5% had a last author affiliated to an African institution. (Figure 2D). There was an equal proportion of non-original (47.8%) and primary research articles (47.1%) published by African first authors (Figure 2B). This was also the case for articles that listed at least one African author. On the other hand, a slightly higher proportion (53.5%) of the articles that listed an African last author were primary research articles (Figure 2C).

**Figure 2:**
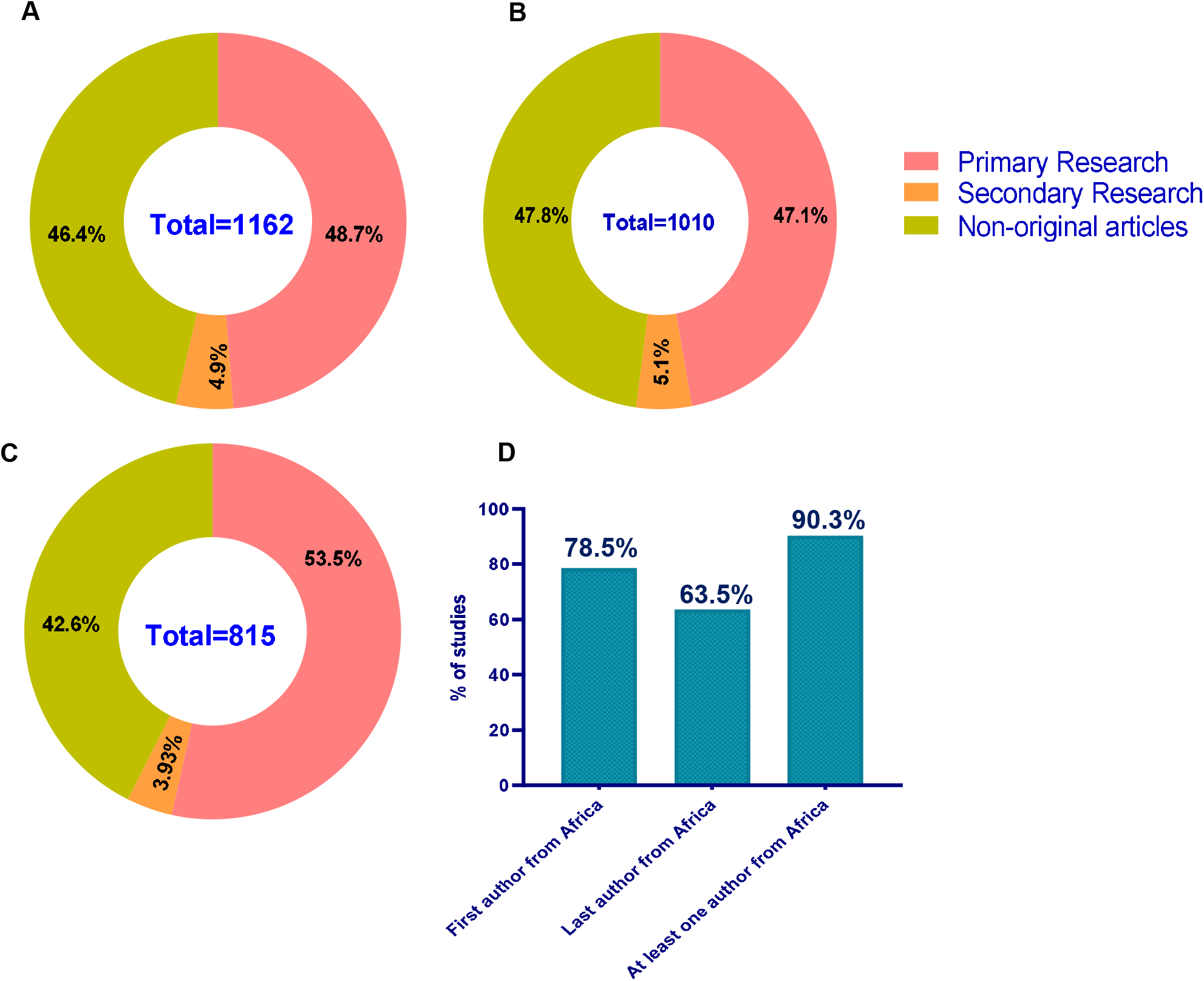
Proportion of articles authored by African researchers and the article type. (A) proportion of research articles that have at least one African author, (B) proportion of research articles that have an African first author, (C) proportion of research articles that have an African last author, and (D) proportion of all articles by author position

### Author Network Analysis and Author Affiliation

A network map of co-authors who contributed to COVID19 research was created at the threshold of 3 documents per author, resulting in 147 collaborating authors (Supplementary figure 1). The top 15 institutions affiliated with the first and last authors are shown (Table 3). Authors from The University of Cape Town (42) and Stellenbosch University (40) published the highest number of articles on COVID-19 in Africa.

**Table 3:** Top 15 first author and last institutions which published COVID-19 articles

| Institutions | Number of first authors |
| --- | --- |
| University of Cape Town, Cape Town, South Africa | 40 |
| Stellenbosch University, Cape Town, South Africa | 39 |
| University of the Witwatersrand, South Africa | 25 |
| University of KwaZulu Natal, South Africa | 25 |
| University of Pretoria, South Africa | 18 |
| Makerere University, Uganda | 22 |
| London School of Hygiene & Tropical Medicine, UK | 12 |
| University of Lagos, Nigeria | 14 |
| University of Tripoli, Libya | 13 |
| KEMRI-Wellcome Trust Research Programme, Kenya | 11 |
| University of Ibadan, Nigeria | 13 |
| Groote Schuur Hospital, South Africa | 9 |
| Mohamed V University, Morocco | 8 |
| Johns Hopkins Bloomberg School of Public Health, USA | 9 |
| University of Zimbabwe, Zimbabwe | 9 |
| Institutions | Number of last authors |
| University of Cape Town, South Africa | 42 |
| Stellenbosch University, South Africa | 36 |
| University of the Witwatersrand, South Africa | 31 |
| University of KwaZulu Natal, South Africa | 22 |
| Makerere University, Uganda | 22 |
| University of Pretoria, South Africa | 19 |
| Cairo University, Egypt | 13 |
| University College London, UK | 13 |
| University of Ibadan, Ibadan, Nigeria | 13 |
| London School of Hygiene & Tropical Medicine, UK | 13 |
| University of Lagos, Nigeria | 12 |
| University of Oxford, UK | 12 |
| University of Tripoli, Libya | 12 |
| Groote Schuur Hospital, South Africa | 8 |
| KEMRI-Wellcome Trust Research Programme, Kenya | 7 |

### Journals and Funding

We ranked the top 15 journals that published the highest number of articles. 174 articles (13.4%) had not yet been peer-reviewed and were retrieved from the pre-print server, MedRxiv. 77 articles (5.9%) were published in the Pan African Medical Journal and 59 (4.5%) were published in the South African Medical Journal (Table 4).

**Table 4:** Top 15 journals in which COVID-19 research in Africa was published.

| Journal | Number of studies |
| --- | --- |
| MedRxiv preprint | 174 |
| Pan African Medical Journal | 77 |
| South African Medical Journal | 59 |
| The American Journal of Tropical Medicine and Hygiene | 31 |
| International Journal of Infectious Diseases | 27 |
| African Journal of Primary Health Care & Family Medicine | 21 |
| Journal of Global health | 22 |
| PLOS one | 18 |
| BMJ Global Health | 15 |
| Lancet Global Health | 12 |
| Clinical Infectious Disease | 12 |
| BMJ | 11 |
| Lancet | 11 |
| Risk Management Healthcare Policy | 11 |
| Travel Medicine and Infectious Disease | 11 |

In addition, we ranked the top 15 funding bodies that provided funding for COVID-19 research in Africa (Table 5). Of the studies that received funding and included information on the funding source, the Wellcome Trust (18 articles) and the Bill & Melinda Gates Foundation (13 articles) were the top 2 funding bodies.

**Table 5:**
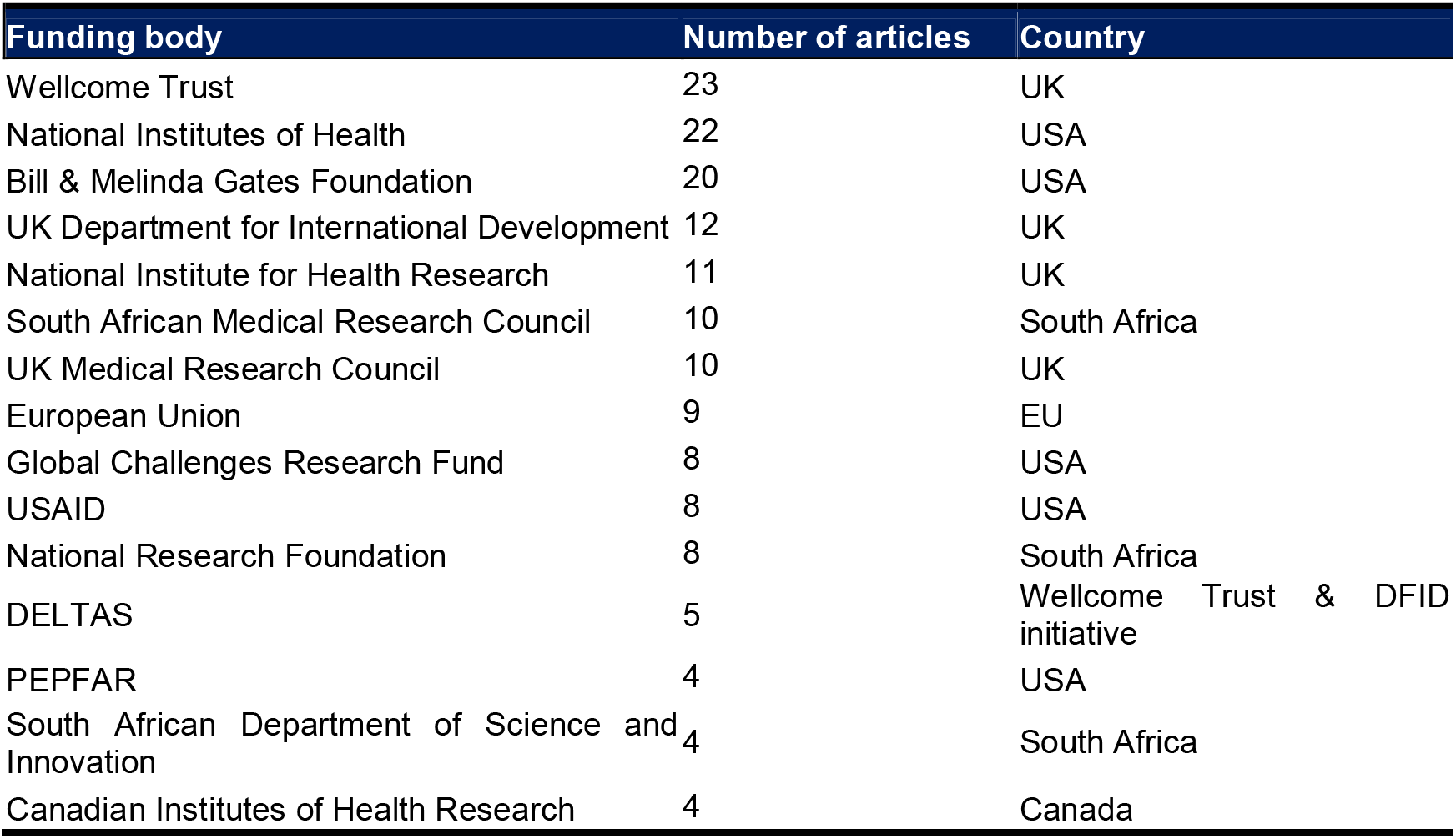
Top 15 funding bodies that provided funding for COVID-19 articles in Africa

### Key Word Analysis and Research Topics

3661 keywords were extracted from the 1296 articles. A density map was generated for keywords with a co-occurrence greater than 5 times thus including 231 keywords on the map. “Humans” was the most frequently used keyword followed by “COVID-19” (Supplementary figure 2). Further, we classified all the articles into specific research groups to determine what the COVID-19 research trends are in Africa and to identify research gaps (Table 6). 25% of the articles had a specific focus on assessing countries’ preparedness and response to the pandemic while 21.6% described the indirect health impacts associated with the pandemic.

**Table 6:** Overview of the top 15 topics covered in COVID-19 research articles in Africa

| Study topic | Number of studies |
| --- | --- |
| Country preparedness and response | 323 (25%) |
| Direct and indirect health impact of pandemic | 282 (21.6%) |
| Transmission | 181 (14%) |
| Knowledge, Attitudes and Practice | 117 (9%) |
| Clinical characteristics | 78 (6%) |
| Socio-economic impacts | 73 (5.6%) |
| Description of pandemic | 47 (3.6%) |
| COVID-19 testing | 30 (2.3%) |
| Genomic sequencing | 19 (1.5%) |
| Role of research | 15 (1.1%) |
| Clinical management | 14 (1%) |
| Ethics | 14 (1%) |
| Therapeutics and vaccines | 13 (1%) |
| Infection prevention and control | 12 (0.9%) |
| Impact of pandemic response | 9 (0.7%) |
| Other | 69 (5.3%) |

Of note, only 1% of the articles focus on therapeutics and vaccines for COVID-19. This is of concern as research that can inform the design of preventative and therapeutic interventions will be important in controlling the pandemic. The least represented research group is virus history and transmission (0.2%). These articles focus on the naming of COVID-19 and the implication of naming viruses by location and a review of the history of viral diseases in light of COVID-19.

## DISCUSSION

As the COVID-19 pandemic evolves in Africa and the rest of the world, efforts are being accelerated to identify effective preventive and therapeutic measures to mitigate its burden. In order to develop these measures, it is important to generate scientific knowledge on the disease and its consequences that is contextually relevant since the pandemic has not evolved uniformly globally. To this effect, there has been an unprecedented volume of COVID-19 research published since the pandemic begun. Africa has contributed to this research and continues to build a body of evidence that can inform local interventions and mitigation strategies. This bibliometric analysis describes the COVID-19 research that has been carried out in Africa.

Among LMICs, nations in sub-Saharan Africa accounts for 10% of the global population and only 1.3% of global health research publications^17^. The analysis of the study designs published showed that almost half (48.6%) of the articles were commentary and editorial pieces and a similar proportion of publications were primary research articles (46.7%). Of the primary research articles identified, 5 (0.004%) were randomized control trials (RCTs) carried out in Africa. RCTs are the gold standard for evaluating effects of healthcare interventions and require substantial skills and experiences. These findings indicate the increase in the number of African researchers and institutions that have the resources, capacity and ability to carry out research using varying study designs depending on the field of interest. On the other hand, there were fewer secondary research articles published (4.6%). However, this is expected as these types of articles require a large volume of primary research which is currently not available as COVID-19 is still a new research area.

Most of the articles included in the analysis focused on the whole continent as the study setting. In the remaining articles, South Africa was the dominant study setting identified. This finding is also in line with other studies that have assessed the volume of all health research from Africa where they report that South Africa has the highest publication output on the continent^17-19^. This is less likely a reflection of prevalence of the disease in the country but other factors such concentration of top research intuitions, availability of local funding, and the country’s gross domestic product (GDP). For example, six of the top 15 institutions contributing to COVID-19 body of research in Africa are based in South Africa and increased capacity for research in terms of number of research institutions is important for increasing productivity. Nachega *et al*. found that the number of public health research institutions present in an African country was associated with research productivity; for every additional research programme, public health research productivity increased by 241%^20^. Secondly, in our analysis of institutions that fund COVID-19 research in Africa, we found that 3 of the top 15 funding bodies were from South Africa, with the other 12 being non-African. Availability of local funding not only increases publication output but is also likely to focus on local issues. Finally, South Africa’s high gross domestic GDP could also account for its high research productivity. Uthman *et al*. reported that an independent factor that influenced research capacity was a country’s GDP^17^. The lower number of articles from most African countries is may be due to a lack of an enabling environment like that in South Africa. In addition, lack of incentives may lead to trained African researchers from certain countries moving to countries with better research infrastructure thereby reducing the number of well-trained scientists on the continent and local research output. Progress towards controlling COVID-19 in Africa will require countries to sustain COVID-19 research and ensure that knowledge generated is also translated into effective policy. This means that African countries should strengthen local research capacity by increasing investment in research and supporting scientists on the continent.

Analysis on the authors of these articles indicate that 90.3% of the articles had at least one African author listed. While this proportion is high, it indicates that 9.7% of articles on Africa and African countries lack African representation. Non-African based researchers often conduct research in Africa on behalf of external agencies in collaboration with African researchers. Although this may be important in the transfer of skills to African researchers, it can also lead to inequity in research partnership arrangements that put African researchers at a disadvantage. The representation and position of authors on publications can be used as a proxy to measure participation and leadership in research. Of the articles that had an African author, 78.5% listed an African first author and 63.5% had an African as last author. Previous studies found that African researchers were underrepresented in studies on infectious diseases in Africa^21^, however, we find that in COVID-19 research, a high proportion of articles were by African authors. This could be due to the urgency of COVID-19 as a public health emergency which has led to a greater demand of research within the continent, increased funding, more time to write and publish, and increased priority for COVID-19 publications^22^. However, a limitation of this analysis is the use of institutional affiliation as an indicator of author nationality. This means that an African author based in a non-African institution would be classified as non-African and a non-African working in an African institution would be classified as African. This misclassification underestimates the contribution of Africa’s large scientific diaspora. Additionally, there was disparity in the geographical representation of African first and last authors. The top four author affiliations were all South African institutions, with the University of Cape Town as the leading institution with the highest number of first and last authors. This is an indicator of the small number of research institutions on the continent and the uneven distribution of these institutions.

Over 13% of articles identified were published in the preprint servers. However, for articles that were peer-reviewed and published in journals, we found that 3 African journals featured in the top 15 journals publishing COVID-19 articles on Africa; Pan African Medical journal, South African Medical Journal and African journal of Primary Health Care & Family medicine. This is promising as availability of local journals where African researchers can publish their work can increase publication output. Most of the research work that is undertaken in Africa is meant for the local audience and addresses local concerns and may not be fairly represented by western publishers. So, it makes sense if such works are published in African journals. However, there is still a long way to go and the development of local journals and publishing houses should be encouraged so as to create a direct avenue for academics and researchers to publish their research findings.

In the articles that declared funding, The Wellcome Trust topped the list of organizations funding COVID-19 research in Africa. Most funding was obtained from organizations in the UK and USA. External funding has greatly contributed to increasing scientific research capacity on the continent. However, this may hinder development of sustainable African-led knowledge production and may deflect research priorities away from local needs. African organizations in the top 15 list of funding bodies were all from South Africa. Few studies were funded by African organizations, and specifically, African governments. This is similar to other previous studies which also observed few funders from the continent^23,24^. Overall, the lack of funding from African governments may reflect lack of economic ability, political will or capacity to fund research. Although increased investment in science by African governments is being made, the continent still falls behind other global regions with African countries spending less than 1% of GDP (on average) on research and experimental development^25^.

Keyword analysis of the articles included in this study found that “COVID-19” and “Human” were the two most occurring (453 times and 417 times respectively) keywords. This is expected as all articles focus on COVID-19 and its effects. This finding is similar to other studies^16,26^. Further, we explored the major themes in COVID-19 research in Africa. Most of the articles (25%) focused on describing the steps that African countries have taken to prepare and respond to the COVID-19 pandemic. Next, 21.6% of articles focused on the direct and indirect health impacts COVID-19 has had by highlighting the disruption and adaptation of health services catering to people with other diseases such as HIV and malaria. Articles describing transmission and clinical characteristics of the virus was another common field of study we identified. However, therapeutics and vaccine studies were underrepresented (1%). Further research efforts in prevention and treatment are both urgent if we are to mitigate the effects of the pandemic.

A major limitation of this study is the inclusion of only English articles. This means that we have missed out on a significant proportion of articles from African countries that do not use English as a main language. In addition, we did not critically appraise the studies/articles included in the analysis and therefore cannot assess the quality of research.

## Conclusion

The COVID-19 pandemic has highlighted the urgent need for research that informs effective action. Africa has contributed to this body of research despite the usual challenges associated with research and development in low resource settings. Contrary to other studies on the output of research publication on infectious diseases, we find that African researchers have played a lead role in publishing COVID-19 research from the continent. However, this productivity is uneven across the continent with only a few countries accounting for majority of the publications. This points to the importance of support for research and development by African governments. Nonetheless, the effort to publish research by Africans for Africans is an indicator of the potential the continent has to contribute to global health research.

## Data Availability

Data will be made available upon request

## Contributors

FHG, SA and EB conceived the protocol; FHG, RO, EK and AM participated in the data collection and extraction; FHG performed data analysis and FHG, SA, RO, EK and EB participated in the article writing.

## Patient consent for publication

Not required.

