## Supplementary File for "A bibliometric analysis of COVID-19 research in Africa"

### Search strategy

The following search terms were used to retrieve studies:

((coronavirus OR "corona virus" OR coronavirinae OR coronaviridae OR betacoronavirus OR covid19 OR "covid 19" OR nCoV OR "CoV 2" OR CoV2 OR sarscov2 OR 2019nCoV OR "novel CoV" OR "wuhan virus") OR ((wuhan OR hubei OR huanan) AND ("severe acute respiratory" OR pneumonia) AND (outbreak)) OR "Coronavirus"[Mesh] OR "Coronavirus Infections"[Mesh] OR "COVID-19" [Supplementary Concept] OR "severe acute respiratory syndrome coronavirus 2" [Supplementary Concept] OR "Betacoronavirus"[Mesh]) AND (("Africa"[MeSH] OR Africa*[tw] OR Algeria[tw] OR Angola[tw] OR Benin[tw] OR Botswana[tw] OR "Burkina Faso"[tw] OR Burundi[tw] OR Cameroon[tw] OR "Canary Islands"[tw] OR "Cape Verde"[tw] OR "Central African Republic"[tw] OR Chad[tw] OR Comoros[tw] OR Congo[tw] OR "Democratic Republic of Congo"[tw] OR Djibouti[tw] OR Egypt[tw] OR "Equatorial Guinea"[tw] OR Eritrea[tw] OR Ethiopia[tw] OR Gabon[tw] OR Gambia[tw] OR Ghana[tw] OR Guinea[tw] OR "Guinea Bissau"[tw] OR "Ivory Coast"[tw] OR "Cote d'Ivoire"[tw] OR Jamahiriya[tw] OR Jamahiryia[tw] OR Kenya[tw] OR Lesotho[tw] OR Liberia[tw] OR Libya[tw] OR Libia[tw] OR Madagascar[tw] OR Malawi[tw] OR Mali[tw] OR Mauritania[tw] OR Mauritius[tw] OR Mayote[tw] OR Morocco[tw] OR Mozambique[tw] OR Mocambique[tw] OR Namibia[tw] OR Niger[tw] OR Nigeria[tw] OR Principe[tw] OR Reunion[tw] OR Rwanda[tw] OR "Sao Tome"[tw] OR Senegal[tw] OR Seychelles[tw] OR "Sierra Leone"[tw] OR Somalia[tw] OR "South Africa"[tw] OR "St Helena"[tw] OR Sudan[tw] OR Swaziland[tw] OR Tanzania[tw] OR Togo[tw] OR Tunisia[tw] OR Uganda[tw] OR "Western Sahara"[tw] OR Zaire[tw] OR Zambia[tw] OR Zimbabwe[tw] OR "Central Africa"[tw] OR "Central African"[tw] OR "West Africa"[tw] OR "West African"[tw] OR "Western Africa"[tw] OR "Western African"[tw] OR "East Africa"[tw] OR "East African"[tw] OR "Eastern Africa"[tw] OR "Eastern African"[tw] OR "North Africa"[tw] OR "North African"[tw] OR "Northern Africa"[tw] OR "Northern African"[tw] OR "South African"[tw] OR "Southern Africa"[tw] OR "Southern African"[tw] OR "sub Saharan Africa"[tw] OR "sub Saharan African"[tw] OR "subSaharan Africa"[tw] OR "subSaharan African"[tw]) NOT ("guinea pig"[tw] OR "guinea pigs"[tw] OR "aspergillus niger"[tw]))

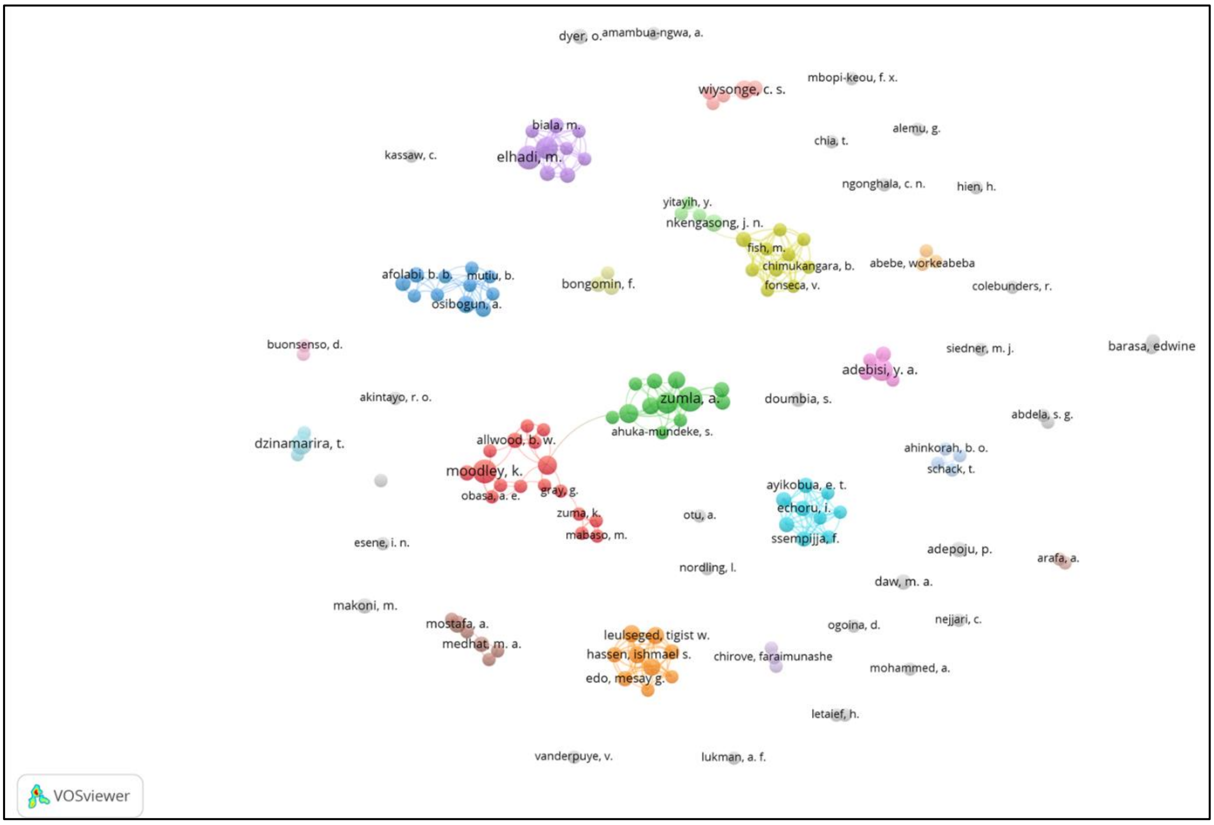


Supplementary figure 1: A network of authors contributing to COVID-19 research in Africa. This includes 147 authors who have contributed not less than 3 papers. Scattered nodes show groups of collaborating authors while links between individual authors within the group are plotted.

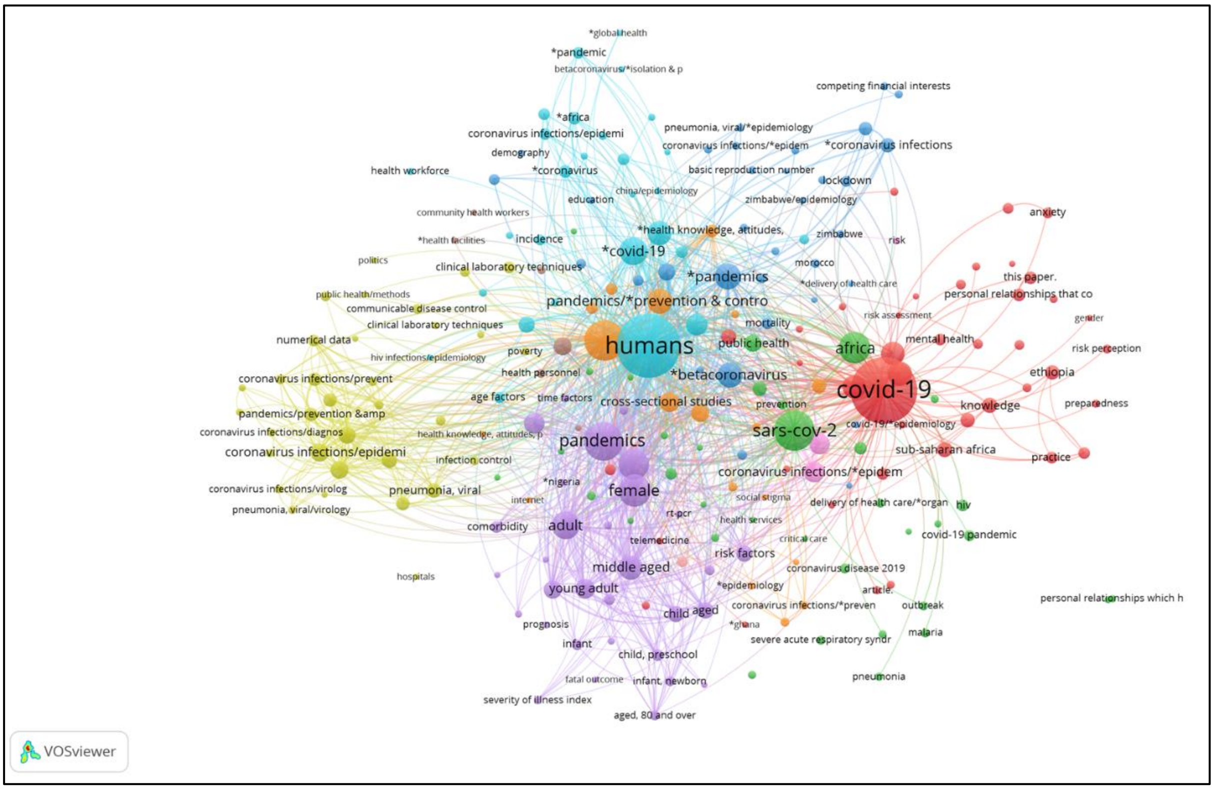


Supplementary figure 2: A network of keywords that co-occur greater than 5 times in the 1296 articles. A total of 231 terms from 3661 keywords were entered into this network and clustered into 6

groups, which are shown with different colors. The most frequent terms are “Humans” (blue) and “COVID-19 (red).
